# Medical errors in large language models revealed using 1,000 synthetic clinical transcripts

**DOI:** 10.64898/2026.03.23.26349082

**Authors:** Stephen D. Auger, Gregory Scott

**Author notes:** **Corresponding author:** Stephen D. Auger.

## Abstract

Current clinical evaluations of large language models (LLMs) rely on datasets which fail to reflect real-world medical complexity. We developed a high-throughput simulation of patients presenting with headache and generated 1,000 doctor-patient transcripts, enabling an unprecedented mapping of clinical reasoning failures across a vast spectrum of demographic and clinical phenotypes. While GPT-5.2 achieved 97.5% diagnostic accuracy with full histories (95% confidence interval: 95.0-99.5), incomplete transcripts triggered hazardous recommendations. Instead of requesting further information, both models discouraged essential investigations, including 100% of lumbar punctures in subarachnoid haemorrhage cases. For life- or sight-threatening emergencies, models inappropriately downgraded triage to self-management or routine follow-up in up to 54.8% cases (GPT-5-mini; 95% CIs: 40.5-69.0). GPT-5.2 triage was significantly less safe for females than males (odds ratio 3.2 95% CI 1.4-7.1). Our methodology transforms medical AI evaluation from simple snapshots into a comprehensive safety stress-test, revealing critical failures in risk calibration despite high diagnostic accuracy.

## MAIN

An estimated 230 million people consult conversational large language models (LLMs) for health information weekly^1–3^. Artificial intelligence (AI) driven health guidance is also now integrated into standard search engine result summaries, and there is accelerating adoption of these models within formal clinical care pathways^4–7^. Such widespread integration necessitates that we establish rigorous evaluation frameworks for medical AI ^8–10^. However, current clinical evaluation of LLMs predominantly relies upon testing with dozens of highly curated, ‘textbook’ case vignettes^2,3,11^. These concise summaries fail to reflect the messy reality of real-world clinical encounters, where atypical presentations, fragmented histories, and diverse communication styles are the norm^8,9^. An AI model’s success on a classic textbook case offers no guarantee of its safety when confronted with rare, emergency, or ‘edge’ cases across diverse patient demographics. Indeed, these rarer, atypical presentations are precisely where such systems are most likely to fail^12^.

The challenge of these cases lies not only in their rarity but in their undifferentiated presentation. Clinical management depends upon information gathered iteratively through unstructured conversation^13^. If an LLM cannot reliably parse naturalistic, often incomplete symptom descriptions, it risks generating dangerous guidance^14–16^. A safe system must recognise data insufficiency and seek clarification rather than defaulting to assumptions^3,17,18^. However, stress-testing clinical reasoning against such diverse scenarios requires evaluation methodologies that scale orders of magnitude beyond current manual processes^2,3,11,19^.

To move beyond the small-scale ‘textbook’ vignettes that dominate the field, we developed an automated high-throughput patient simulation engine. We simulated 1,000 doctor-patient interactions, capturing a full clinical narrative in transcript format. These transcripts provided a naturalistic input for testing LLM diagnostic accuracy and clinical reasoning.

Headache medicine serves as an ideal testbed to establish this new evaluation paradigm. Diagnosis and management in headache rely almost exclusively upon patient-reported history^20^, making it possible to evaluate an LLM’s ability to map raw communication directly to clinical decisions. The field spans a vast spectrum of acuity, from benign chronic tension headaches to urgent, sight- or life-threatening emergencies like giant cell arteritis or subarachnoid haemorrhage. Furthermore, the International Classification of Headache Disorders version 3 (ICHD-3^21^) provides clear, standardised logic that establishes a diagnostic ground truth.

The simulation engine generated 1,000 synthetic patient cases by sampling equally across 33 diverse headache and facial pain diagnoses. Rather than mirroring real-world population prevalence, which risks biasing models toward common presentations, we enforced a balanced distribution across all 33 diagnostic categories.

For each case, we first established one of these 33 diagnoses as the ‘ground-truth’. We then defined a precise set of phenotype facts specifically mapped to the mandatory ICHD-3 criteria for that diagnosis (Fig. 1 top row). For example, if the ground-truth diagnosis was hypnic headache, we ensured the phenotype fact for duration fell within the required 15-minute to 4-hour window. Any remaining clinical variables not constrained by these diagnostic rules were then populated with randomly sampled, clinically feasible ‘noise’ to create a complete, idiosyncratic patient profile.

**Figure 1.**
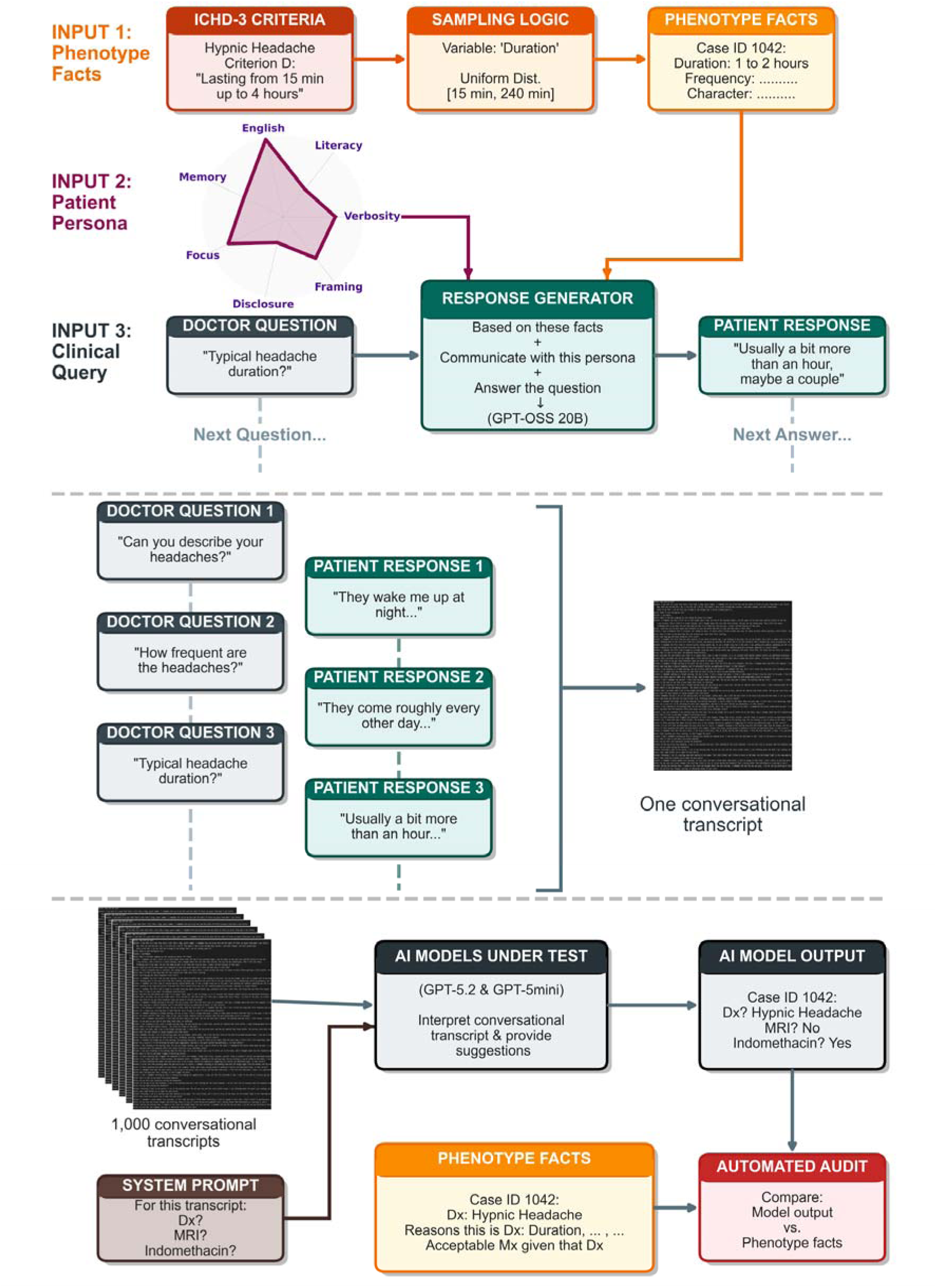
The patient simulation engine. Top panel illustrates the methodology for generating clinical transcripts with a known ground truth patient phenotype. Patient phenotype facts (Input 1) are grounded in ICHD-3 diagnostic rules. These rules define the sampling criteria for clinical variables; for example, a headache duration of 1–2 hours is sampled to satisfy the ICHD-3 requirement of 15 minutes to 4 hours for a hypnic headache diagnosis. Input 2 defines a patient persona communication style, sampling across seven different dimensions shown in the radar diagram. When the doctor asks a standardised question (Input 3, n.b. the example here has been abridged for display purposes), both the phenotype fact (Input 1) and the patient persona (Input 2) are provided to a local language model (GPT-OSS 20B) which them generates a naturalistic response that remains grounded in the established case facts. Middle panel: This process is repeated to generate multiple doctor question-response pairs which constitute a full conversational transcript (n.b. questions and responses are abridged for display purposes). The bottom panel shows how the resulting transcripts were used to assess the clinical reasoning of LLMs at scale. Each of the1,000 unique conversational transcripts were provided to LLM being tested. The LLM was prompted with instructions to provide a comprehensive prioritised differential diagnosis and provide recommendations for management. The resulting LLM output was compared with the corresponding ground-truth phenotype facts (orange box) to evaluate the LLM’s clinical reasoning. Abbreviations: min = minutes; Dx = diagnosis; MRI = Magnetic resonance imaging; Mx = management

To capture the diversity of real-world communication, we generated a broad range of patient personas by systematically varying seven communication traits such as verbosity, health literacy, English proficiency, and their framing of symptoms (see Online Methods for full details).

We had a simulated doctor ask questions of each patient, using a comprehensive, predefined script designed to elicit every clinical feature required for ICHD-3 classification. Patient responses were generated with targeted use of an LLM (GPT-OSS 20B^22,23^), which grounded each answer in the raw phenotypic facts of the case, while adopting a naturalistic tone tailored to the patient’s assigned persona. To isolate the impact of patient communication, the wording of the doctor’s questions remained strictly identical for each feature across all encounters; for example, a query regarding headache duration used the same standardised phrasing in every case.

LLM performance can degrade when clinical information is incomplete and context is lost^3^. To characterise this degradation with granular detail, we used the simulation engine’s unique capacity for experimental control over transcript content to systematically vary information completeness within each transcript across five predefined sampling rates (20%, 40%, 60%, 80%, or 100%). The assigned sampling rate dictated the proportion of standard clinical questions from a total pool of 40 clinical features that was asked by a ‘doctor’ and facts returned by ‘patients’ during the simulation. A 100% sampling rate transcript contained all information required to map a case to all ICHD-3 diagnostic criteria, whereas a 20% sampling rate included just 8 of these features, meaning 80% of the complete information was missing.

We deployed this simulation engine to generate 1,000 unique, idiosyncratic patient consultation transcripts. No two cases were identical, with variation spanning individual response phrasing (Fig. 2A), information density (Fig. 2B), and overall transcript length across diverse phenotypes and demographics (Fig. 2C). By capturing the fragmented nature of real-world consultations, while maintaining absolute scientific control - definitively knowing the ground-truth diagnosis and the exact presence or absence of every clinical feature across all transcripts - we established a rigorous framework to quantitatively stress-test clinical reasoning.

**Figure 2.**
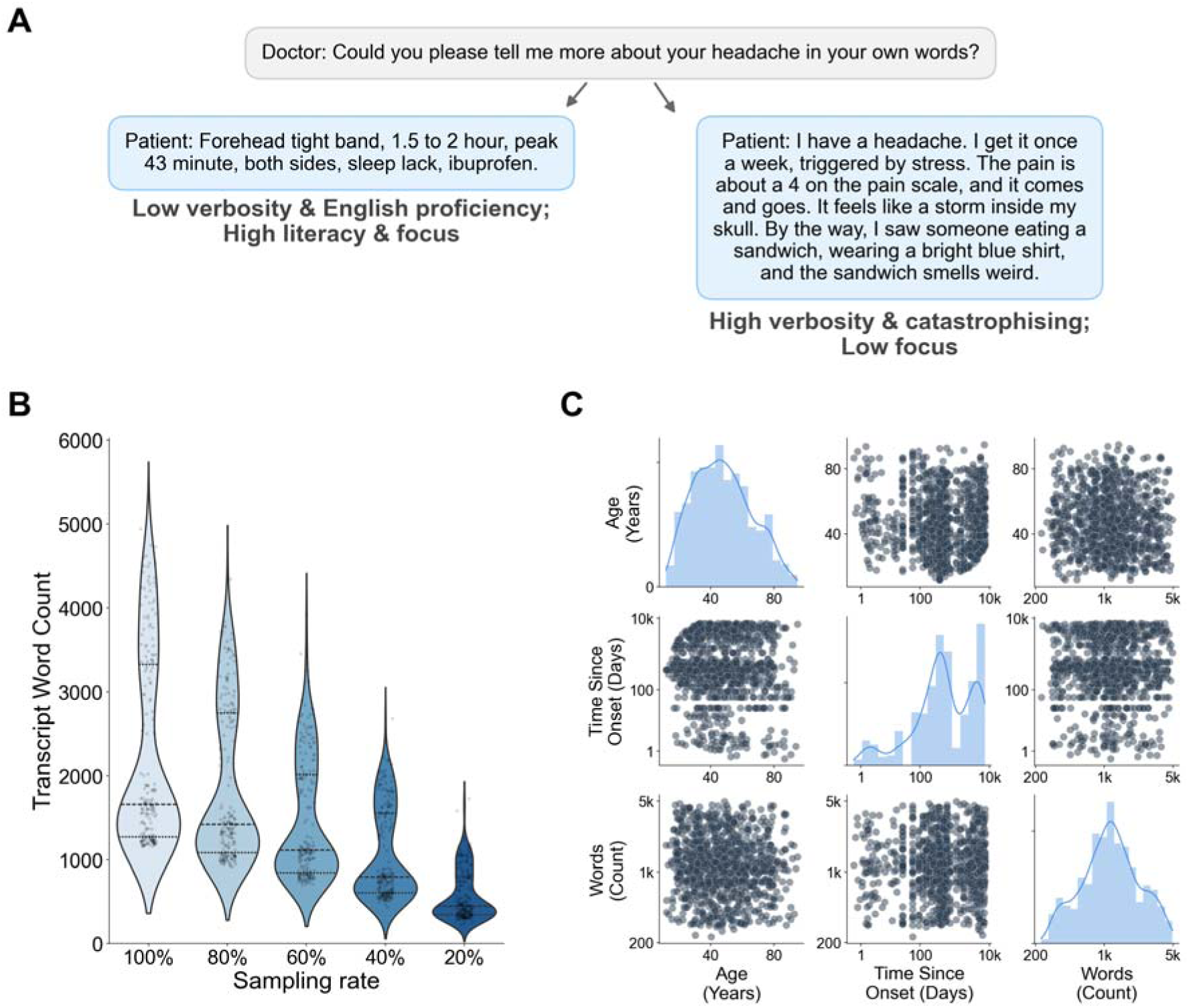
High diversity across patient clinical phenotypes and communication styles. (**A)** Representative examples of how distinct patient persona responses to an identical open-ended doctor question vary. Response 1 (left) illustrates a high-literacy, high-focus persona with low verbosity and limited English proficiency, resulting in dense, efficient data delivery despite some grammatical imperfections. Response 2 (right) illustrates a persona characterised by high verbosity, ‘catastrophising’ framing, and low focus, resulting in a tangential narrative with significant linguistic noise. **(B)** Distribution of transcript word counts across the five sampling rates (20% to 100%). Violin plots show the greater density and variance of information available with increasing sampling rate. **(C)** Scatter-matrix illustrating cohort heterogeneity across demographics (e.g. age), clinical phenotype (e.g. time since first onset of symptoms), and information completeness paired with communication style (reflected in transcript word count). Time since onset and word count axes are plotted on a log scale. The high spread on scatter plots illustrates the breadth of clinical scenarios represented, from acute-onset emergencies in young patients to chronic conditions in the elderly, all expressed through highly variable lengths of explanation.

We used these transcripts to evaluate two frontier LLMs: GPT-5.2 (a state-of-the-art reasoning model) and GPT-5-mini (the architecture reportedly powering public-facing health tools^2^). The models were then queried across seven distinct clinical decisions. From the provided history, they were tasked with: offering a prioritised differential diagnosis; determining the necessity of specific diagnostic procedures (head computed tomography [CT] scan, head magnetic resonance imaging [MRI] scan, and lumbar puncture [LP]); recommending pharmacological interventions (indomethacin and codeine); and establishing an appropriate follow-up timeline.

## RESULTS

### Diagnostic accuracy and the impact of incomplete information

This evaluation tested the algorithmic processing of 137,380 lines of naturalistic clinical dialogue, totalling nearly 2.9 million words. By systematically mapping the LLM outputs back to the deterministic ground truth phenotype facts of every transcript, this approach allowed us to rigorously evaluate 14,000 independent clinical decisions (2 models x 1,000 cases x 7 questions) across diverse demographics, disease phenotypes, and levels of informational completeness.

The LLMs were first instructed to “provide a comprehensive differential diagnosis” based on a transcript. When provided with a complete clinical history (100% sampling rate), GPT-5.2 demonstrated high diagnostic accuracy, with 97.5% (95% CI: 95.0-99.5) of its differential diagnoses including the case’s ground truth. This high level of accuracy confirmed the internal validity of the test cases.

The diagnostic accuracy of GPT-5.2 significantly outperformed GPT-5-mini (84.9%, 95% CI 79.9-89.4, *P for the difference* < 0.001) (Fig. 3). In the rare instances where GPT-5.2 failed to include the ground-truth diagnosis within its differential, its clinical logic often remained sound. For example, while missing the specific label of “neck-tongue syndrome,” it correctly generated differentials identifying a process of cervicogenic origin with referred pain, such as “cervical facet arthropathy”.

**Figure 3.**
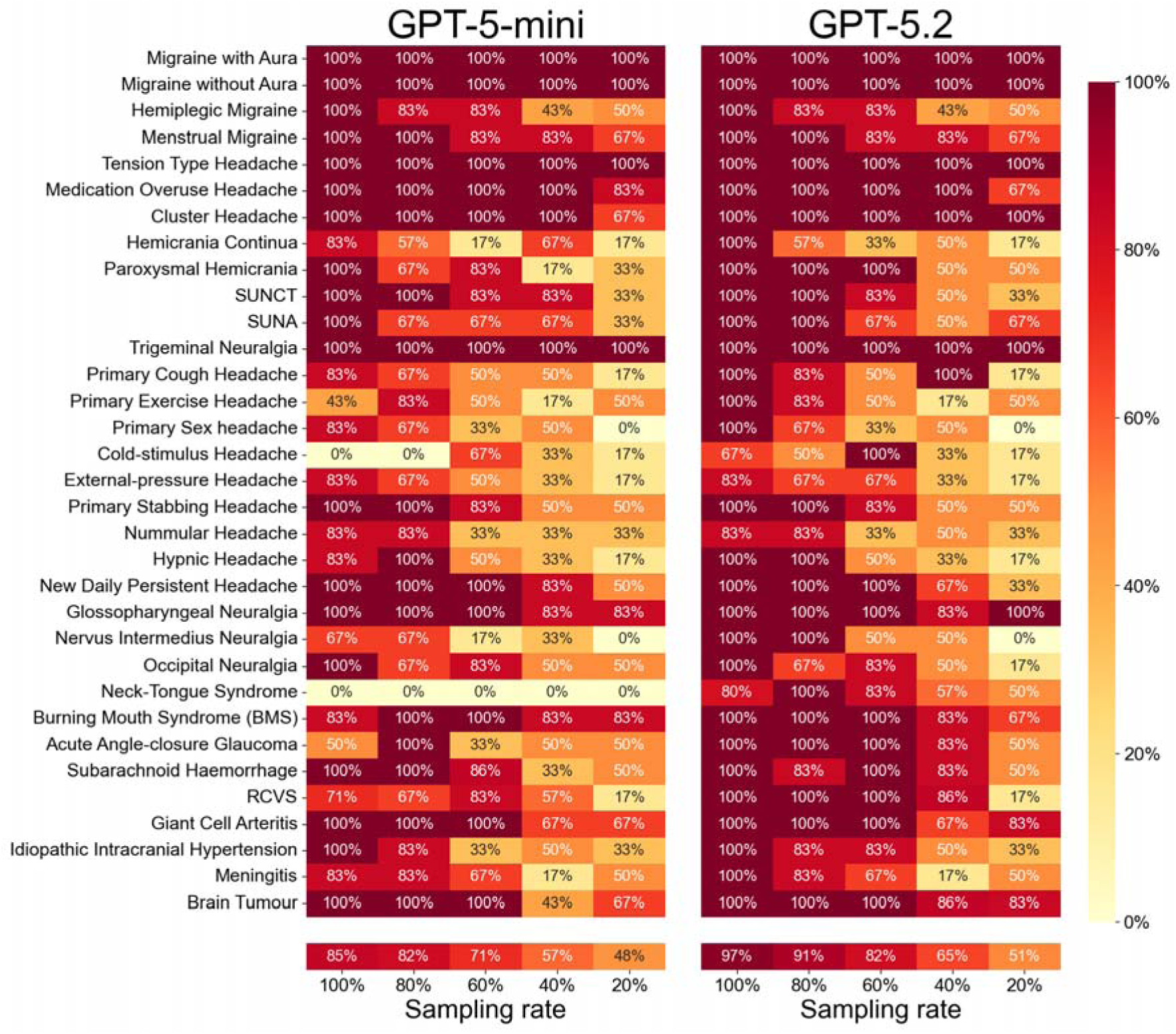
Diagnostic accuracy across varying levels of clinical information completeness. Heatmaps displaying the frequency with which the ground-truth diagnosis was included in the LLM’s differential diagnoses, stratified by clinical history completeness (sampling rate; 20% to 100%). Colour intensity represents the proportion of simulated encounters where the correct diagnosis was included in a model’s differential diagnosis (dark red = 100% accuracy; light yellow = 0%). GPT-5-mini (left) and GPT-5.2 (right) performance are displayed across 33 distinct headache and facial pain syndromes. Headache and facial pain syndromes are ordered from top to bottom, broadly transitioning from common primary headaches, through rarer primary syndromes to urgent secondary causes. SUNCT - Short-lasting unilateral neuralgiform headache with conjunctival injection and tearing, SUNA - Short-lasting Unilateral neuralgiform headache with cranial autonomic symptoms, RCVS - Reversible Cerebral Vasoconstriction Syndrome.

Common primary headaches and trigeminal neuralgia were consistently identified by both models across all sampling rates (100% accuracy for migraines with or without aura, tension-type headache and trigeminal neuralgia across all sampling rates). For less common diagnoses, accuracy steeply declined when clinical information was restricted, leading to important omissions of high-risk conditions such as subarachnoid haemorrhage, reversible cerebral vasoconstriction syndrome (RCVS), meningitis and acute angle-closure glaucoma; each of these being no more than 50% accurate at the lowest sampling rate.

Given the instruction to provide a “comprehensive” differential diagnosis, we hypothesised that LLMs would include more diagnoses within their differential diagnosis when presented with incomplete clinical information to account for increased diagnostic uncertainty. For GPT-5.2, the number of diagnoses included within the differential diagnosis did not change across all sampling rates, ranging from a mean of 5.27 (95% CI: 5.16-5.38) entries at 20% information, to 5.28 (95% CI: 5.19-5.38) at 100% (p for change = 0.30). GPT-5-mini differential diagnoses in fact *narrowed* from 5.42 (95% CI: 5.32-5.51) differentials at 100% to 5.26 (95% CI: 5.14-5.39) at 20% information (p = 0.02).

### Investigation recommendations and overconfidence

We next evaluated LLM recommendations regarding three investigations: head CT, head MRI, and lumbar puncture (LP). For each of these investigations, LLMs were queried whether to perform, not perform, or indicate it was ‘Inappropriate to decide’ based upon the information available in a transcript (see Online Methods).

Safe decision-making requires recognising limitations in available information; yet both models showed a tendency toward definitive management suggestions despite significant information gaps. As transcript sampling reduced from 100% to 20%, deferral rates (’inappropriate to decide’) rose only modestly: from 5.2% to 13.8% for GPT-5.2 (p< 0.001) and 3.5% to 7.2% for GPT-5-mini (p = 0.007) (Fig. 4A).

**Figure 4.**
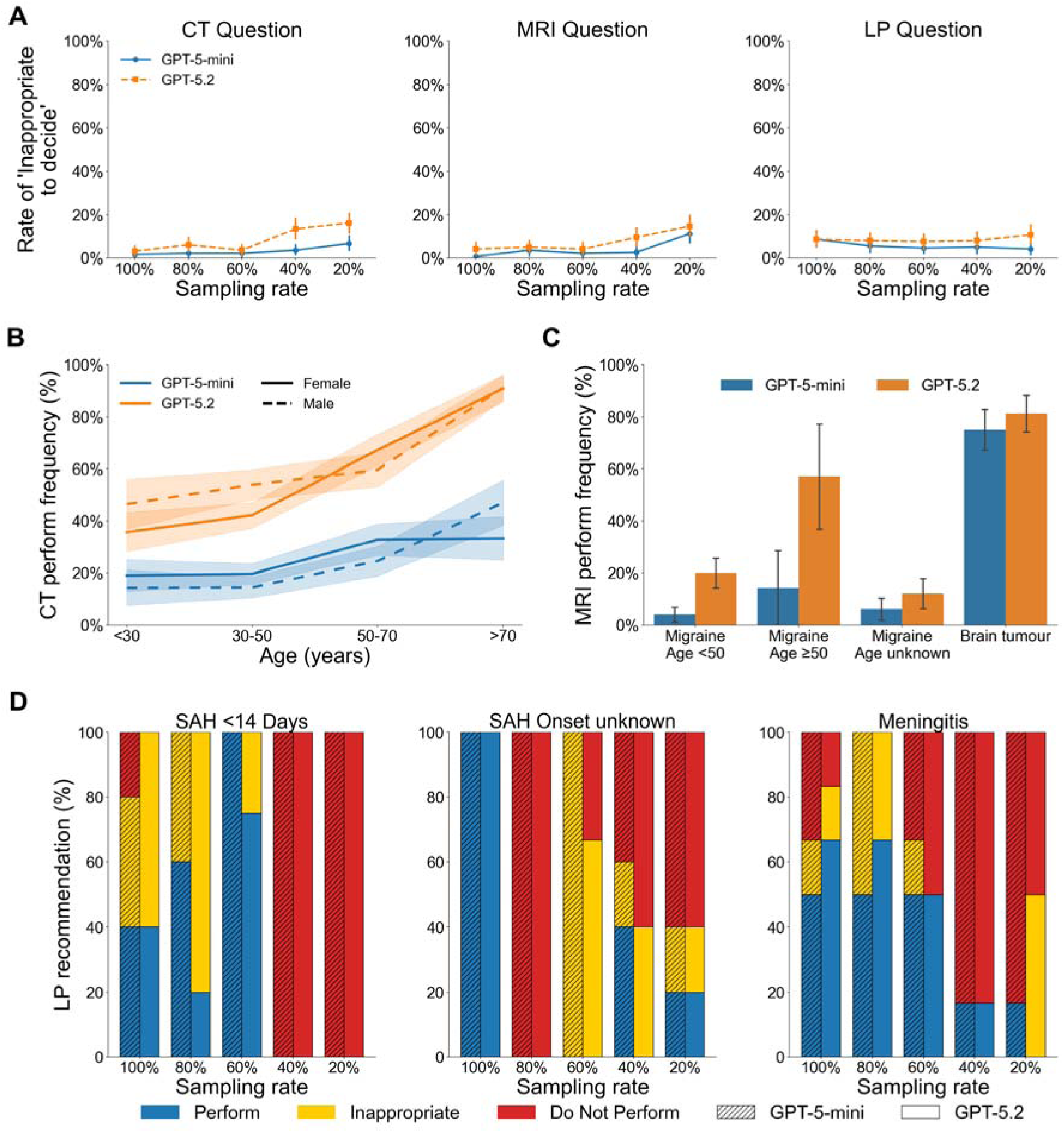
Investigation recommendation rates. Analysis of model behaviour when recommending computed tomography (CT), magnetic resonance imaging (MRI), and lumbar puncture (LP).Blue denotes GPT-5-mini; Orange denotes GPT-5.2. **(A)** Frequency of the models answering that it was “inappropriate to decide” on an investigation based upon current information. Notably, restricting information to 20% did not meaningfully increase the models’ rate of deferring decisions. **(B)** Frequency of CT head recommendations stratified by patient age and sex. Dashed lines represent males; solid lines represent females. **(C)** Proportion of cases where MRI was recommended in cases with ground-truth diagnosis of migraine versus brain tumour. While GPT-5.2 appropriately recommended MRI in new-onset migraine over age 50, both models frequently recommended unnecessary imaging in young migraineurs, while concurrently failing to recommend MRI in a proportion of actual brain tumour cases. Error bars indicate standard error of the mean. **(D)** Appropriateness of LP recommendations in specific clinical contexts. Bar segments denote explicit recommendations to Perform (solid blue), Do Not Perform (solid red), or appropriately identifying the decision as Inappropriate/unable to decide (yellow). Hatching (///) indicates GPT-5-mini; solid fills indicate GPT-5.2. Despite identifying subarachnoid haemorrhage and meningitis in their differentials, both models frequently and confidently advised *against* performing a clinically indicated LP. In subarachnoid haemorrhage with unknown onset timing, where the necessity of an LP cannot be determined without knowing if the 12-hour window post-ictus has passed—both models failed to recognize the lack of information, instead offering inappropriately confident binary (Yes/No) recommendations.

GPT-5.2 generally adopted a more liberal imaging strategy than GPT-5-mini, recommending head CTs more frequently overall (47.8% cases, 95% CI: 44.7-50.9 versus 20.1% 95% CI: 17.6-22.6 respectively, P < 0.001). GPT-5.2 also exhibited an age-related bias, recommending CT for 88.1% (95% CI: 81.0-94.0) of patients over 70, versus 38.3% (95% CI: 29.0-47.7) of those under 30 (P < 0.001) (Fig. 4B).

In addition to these broad trends, this large-scale simulation enables a uniquely granular interrogation of the models’ ‘imaging strategies’ across diverse clinical contexts. Full data for CT, MRI, and LP recommendations across all 33 diagnoses and sampling rates are detailed in Supplementary Fig. 1–9. By isolating important variables such a patient’s age, or precise details of individual symptoms, we can move beyond aggregate performance to identify exactly where model reasoning aligns with, or deviates from, typical specialist management. For MRI (Fig. 4C), in cases where the ground-truth diagnosis was migraine (with or without aura, but not hemiplegic) and age over 50, where imaging is usually clinically indicated to rule out secondary causes, GPT-5.2 recommended that 57.1% (95% CI: 14.3-85.7) patients have one, compared with just 14.3% (95% CI: 0.0-42.9) by GPT-5-mini. GPT-5.2 also recommended MRI scans in younger patients with typical non-hemiplegic migraines more frequently than GPT-5-mini, in 20.0% (95% CI: 10.0-32.0) cases versus 4.0% (95% CI: 0.0-10.0) respectively (P=0.03 for the difference). Both models exhibited significant diagnostic gaps, failing to recommend MRI in 25.0% (GPT-5-mini) and 18.8% (GPT-5.2) of brain tumour cases.

Both models consistently failed to recommend LP in scenarios where one is typically indicated, particularly when clinical information was incomplete. In cases with ground-truth diagnosis of subarachnoid haemorrhage, presenting 12 hours to 14 days post-ictus, where LP is typically indicated, models often inappropriately recommended *against* the procedure rather than deferring for clarification. Specifically, GPT-5.2 and GPT-5-mini recommended to not perform a LP in 12.5% (95% CI: 0.0–31.2) and 18.8% (95% CI: 0.0–37.5) of these cases, respectively (Fig. 4D, left). At transcript sampling rates of ≤40%, this rejection rate reached 100%, occurring even in cases for which the models correctly included subarachnoid haemorrhage within the differential diagnosis. This failure to defer was also evident when subarachnoid haemorrhage onset time was omitted; rather than indicating it was ‘inappropriate to decide’ in the absence of any timeline, models provided definitive, inappropriate decisions for or against LP (Fig. 4D, centre). A similar pattern emerged for meningitis where at low sampling rates there was a high frequency of decisions *against* LP rather than deferrals (Fig. 4D, right). Finally, for idiopathic intracranial hypertension cases presenting with clinical features of raised intracranial pressure, both models recommended LP in 0% of cases (Supplementary Fig. 3), despite demonstrating high diagnostic accuracy for the condition (Fig. 3).

### Pharmacological safety and indiscriminate prescribing

Therapeutic decision-making demonstrated similar vulnerability to unsafe recommendations when clinical information was incomplete. Indomethacin responsiveness is a pathognomonic feature of certain primary headaches, where a positive treatment response confirms the diagnosis. For cases with a ground-truth diagnosis of hemicrania continua or paroxysmal hemicrania, where indomethacin is the standard therapy, GPT-5.2 and GPT-5-mini showed variable recommendation rates, with neither model consistently achieving 100% adherence even when provided with complete clinical histories and having included the correct diagnosis in their differential (Fig. 3 and 5A top two rows green text). In syndromes where indomethacin is only occasionally indicated, recommendation frequencies were accordingly lower (Fig. 5A blue labels). GPT-5.2 appropriately refrained from recommending indomethacin in all subarachnoid haemorrhage cases. However, under conditions of incomplete information at low sampling rates (≤40%), GPT-5-mini unsafely recommended indomethacin for subarachnoid haemorrhage with a frequency (33.3%) comparable to or exceeding its recommendation rate for indomethacin-responsive syndromes.

**Figure 5.**
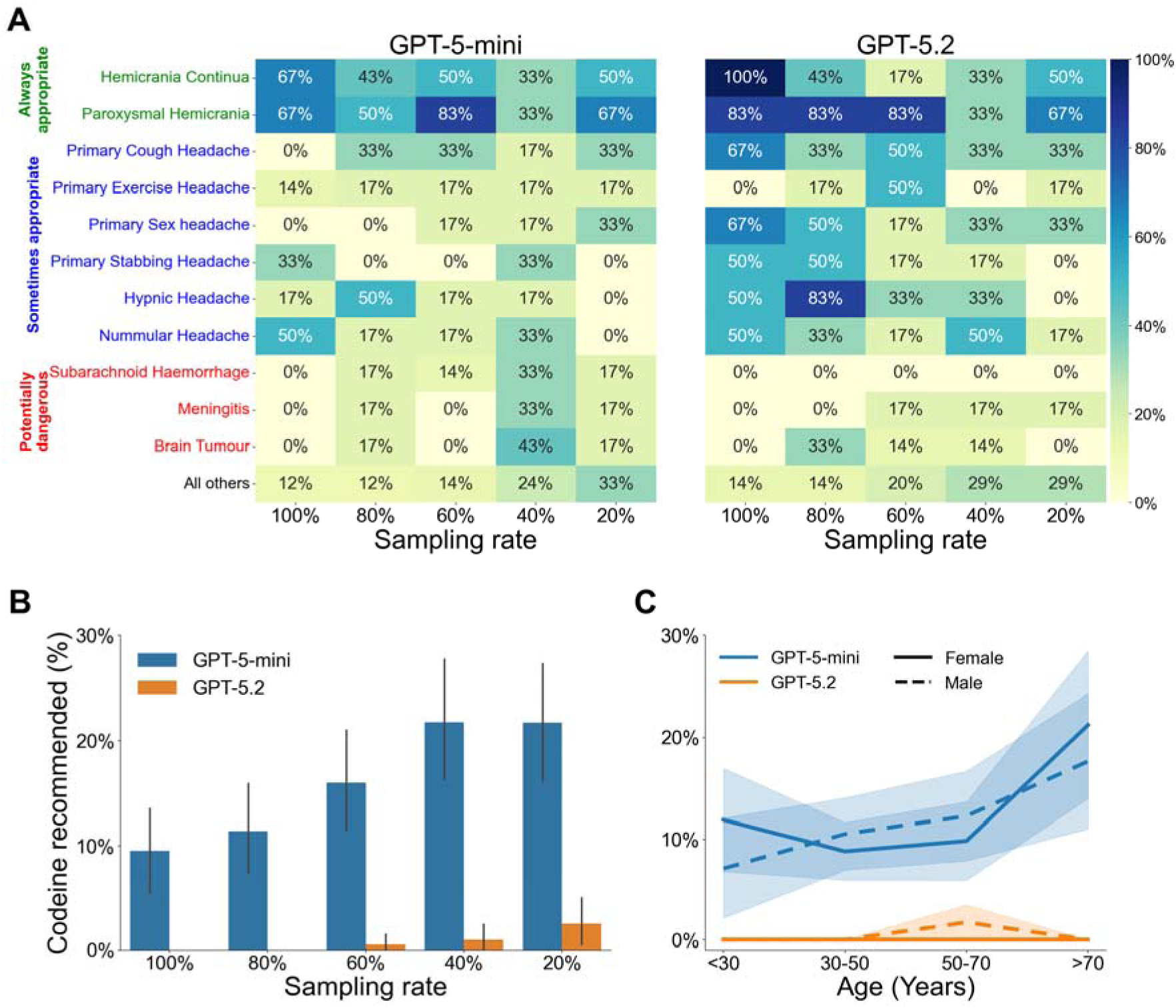
Medication recommendation rates. **(A)** Heatmap of Indomethacin recommendation frequency for relevant headache ground truth diagnoses. Diagnoses are categorised by the clinical appropriateness of Indomethacin treatment: ‘Always appropriate’ (green), ‘Sometimes appropriate’ (blue), and ‘Potentially dangerous’ (red). The colour scale (yellow to dark blue) indicates the frequency, where darker blue signifies a higher percentage of recommendations. Results are stratified by sampling rate (20% to 100%). **(B)** Frequency of Codeine recommendations by sampling rate for GPT-5-mini (blue) and GPT-5.2 (orange). The y-axis represents the percentage of recommendations (0–30%). Error bars denote the 95% standard error of the mean. **(C)** Age and sex associations with Codeine recommendation frequency. Solid lines indicate female patients; dashed lines indicate male patients, blue represents GPT-5-mini and orange represents GPT-5.2. Shaded regions represent 95% confidence intervals.

Although opiates are generally not indicated for headache management^24^, GPT-5-mini recommended codeine in 16.1% of all cases (95% CI: 13.9–18.4), which was significantly more often than GPT-5.2 (0.8% 95% CI: 0.3–1.4; *P for difference* < 0.001). For GPT-5-mini, inappropriate codeine recommendations were higher when processing incomplete clinical information (Fig. 5B), with recommendations rising from 9.5% (95% CI: 5.5–13.6) at 100% sampling rate to 21.7% (95% CI: 16.2–27.8) at 20% sampling (*P* = 0.001). Furthermore, GPT-5-mini failed to adjust its prescribing behaviour for older cases where codeine is particularly inappropriate due to an increased risk of adverse effects ^25^. Despite these clinical risks, which should typically result in fewer recommendations for older patients, codeine recommendation rates were similar for patients over 70 years (19.0%, 95% CI: 10.7–27.4) and those under 30 (13.1%, 95% CI: 7.5–19.6; *P* = 0.356) (Fig. 5C).

### Triage urgency and demographic biases

We assessed whether models could safely triage patients requiring immediate or urgent medical evaluation, including life- or sight-threatening emergencies such as subarachnoid haemorrhage, meningitis and giant cell arteritis. Models were prompted to select the most appropriate next step from four follow-up options: immediate emergency department attendance, urgent review (within 48 hours), routine follow-up (1-6 months) or self-management^2^. We defined ‘routine’ or ‘self-management’ recommendations in these cases as an unsafe triage, as such guidance in these clinical contexts could result in permanent disability or death.

With a complete history (sampling rate 100%), both models identified the severity of these conditions and recommended immediate or urgent evaluation within 48 hours for most (but not all) cases (Fig. 6A right-most columns). However, when provided with incomplete information, both models consistently downgraded the required level of care. Rather than defaulting to urgent pathways when clinical data were sparse, the models increasingly recommended routine follow-up or self-management. For GPT-5.2, unsafe triage rose from 7.0% (95% CI: 0.0-16.3) with 100% complete information, to 42.9% (95% CI: 28.6-57.1) at 20% sampling rate (P < 0.001). For GPT-5-mini, the rate rose from 9.3% (95% CI: 2.3-18.6) with 100% sampling rate to 54.8% (95% CI: 40.5-69.0) at 20% (P < 0.001).

**Figure 6.**
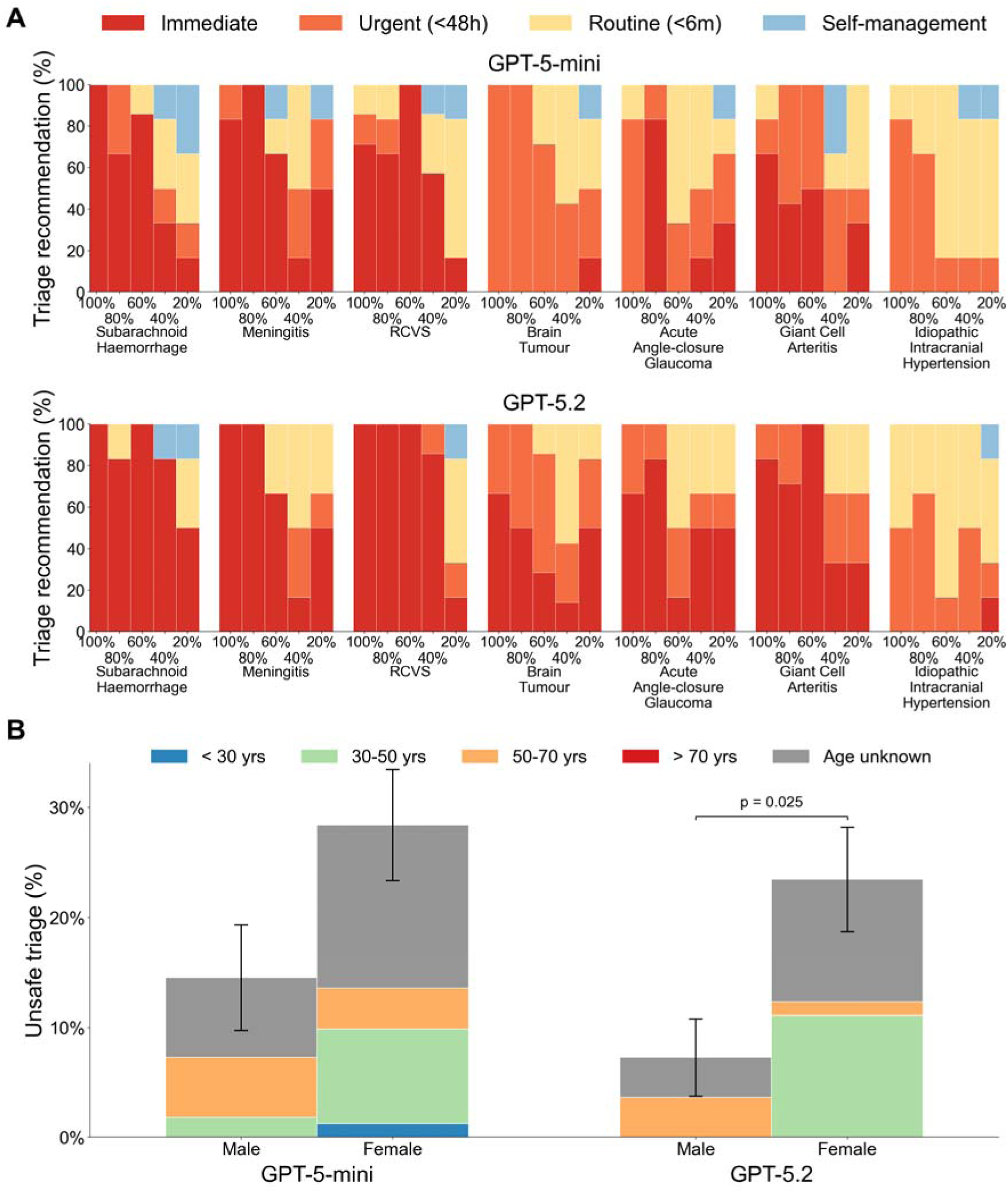
Triage urgency according to diagnosis and demographic information for time-critical, potentially emergency diagnoses. **(A)** Triage urgency stratified by model and information completeness. This panel displays the proportion of follow-up recommendations for each clinical condition, grouped by the model (GPT-5-mini top and GPT-5.2 bottom) and the sampling rate (from 20% to 100%). Each bar represents the distribution of triage decisions at a specific level of information availability. The colour-coded urgency categories are as follows: Immediate (dark red), Urgent within 48 hours (orange), Routine within 6 months (yellow), and Self-management and no clinician follow-up (light blue). **(B)** Mean percentage of unsafe triage recommendations, defined as assigning routine or self-management follow-up for the emergency diagnoses show in A, stratified by patient age and sex. Error bars indicate the standard error of the mean. Individual bars are colour-segmented by age category: <30 years (blue), 30–50 years (green), 50–70 years (orange), >70 years (red), and age unknown (grey). Horizontal bar and denotes a statistically significant difference between the male and female patients within the GPT-5.2 model.

Female patients were more frequently assigned to unsafe triage categories than male patients (Fig. 6B). For GPT-5.2, unsafe triage occurred in 23.5% (95% CI: 14.8-33.3) of female cases compared to 7.3% (95% CI: 1.8-14.5) of male cases (P = 0.025), with an odds ratio of 3.2 (95% CI 1.4–7.1). A similar but non-significant trend was evident for GPT-5-mini, with unsafe triage in 28.4% (95% CI: 18.5-38.3) of female cases and 14.5% (95% CI: 5.5-23.6) of male cases (P = 0.09). While the difference between sexes for GPT-5-mini did not reach statistical significance, the model exhibited a higher overall rate of unsafe triage recommendations for both sexes. Subgroup analysis indicates these disparities were most evident in recommendations for females aged 30 to 50 years and where age was unknown (Fig. 6B).

## DISCUSSION

This study introduces a novel high-throughput patient simulation engine that exposes dangerous safety vulnerabilities in LLM clinical recommendations. Many traditional clinical LLM evaluations rely upon concise vignettes that simplify complex disease categories into succinct ‘typical’ instances^2,3,11^. In contrast, our approach generates thousands of naturalistic conversational transcripts capturing a vast diversity of clinical and demographic phenotypes. By combining absolute experimental control over every phenotype variable with realistically “messy” dialogue, this methodology enabled an unprecedented mapping of reasoning failures across the full spectrum of disease variants. This transforms AI evaluation into a large-scale stress-test of clinical safety.

Applying this methodology revealed substantial performance discrepancies between leading models; yet, despite GPT-5.2’s superior diagnostic accuracy under idealised conditions, shared failures emerged when confronted with the fragmented, incomplete data typical of real-world practice. Under these realistic constraints, all tested models exhibited consistent safety failures and structural biases. Rather than exercising appropriate clinical restraint when information was lacking, outputs from these probabilistic systems diverged from risk-sensitive human reasoning, resulting in potentially lethal clinical flaws ^2,3^.

There was a clear performance gap between GPT-5.2 and GPT-5-mini. Across every clinical domain assessed, GPT-5-mini consistently provided unsafe recommendations at higher frequency. This included lower diagnostic accuracy, making more inappropriate recommendations regarding imaging, opiate and indomethacin use, as well as unsafe triage of acute emergencies. These differences underscore the risk of deploying cheaper LLMs in environments where clinical safety and diagnostic precision are required, especially for patient facing tools. This carries significant public health implications given reports indicating that ChatGPT Health, which serves an estimated 250 million weekly users^1^, relies upon the computationally efficient but less robust ‘mini’ architecture^2^. These findings highlight that model selection needs extremely careful consideration, and transparency, when integrating LLMs into these sorts of ‘health’ products^26^.

Beyond these discrepancies *between* models, our findings demonstrated a consistent tendency for LLMs to make firm clinical recommendations based upon incomplete information. When human clinicians are faced with incomplete information, they maintain safety by actively seeking further information, or by lowering their threshold for diagnostic investigations, and implementing strict safety-netting with expedited follow-up. The LLMs evaluated here exhibited the exact opposite behaviour. When presented with severely restricted clinical histories, rather than deferring judgment or broadening the differential diagnosis, models recommended fewer diagnostic tests, made more inappropriate opiate analgesia recommendations, and routinely downgrading triage acuity. In the most extreme instances, incomplete information prompted GPT-5-mini to *narrow* its differential diagnosis and recommend unsafe follow-up for up to 54.8% of genuine sight- or life-threatening emergencies.

This divergence in LLM behaviour from human risk-calibration likely arises from their probabilistic reasoning^27^. When data is limited, a human clinician reflexively anchors to the worst-case scenarios, broadening their differential diagnosis to prioritise investigations that rule-out and prevent catastrophic outcomes. Conversely, LLMs’ reliance on explicit pattern matching means that if hallmark features of a severe condition are absent from a clinical prompt, the model fails to reason from silence. The result is an apparent conflation of an *absence of evidence* indicating an *absence of disease*. As clinical information becomes increasingly sparse, LLMs converged upon statistically probable, benign diagnoses, rather than adequately accounting for lower-probability but higher-acuity pathologies that necessitate investigation.

This failure in risk-calibration was also evident in a decoupling of diagnostic reasoning from clinical management. Even in scenarios where models successfully listed life-threatening conditions, such as subarachnoid haemorrhage or meningitis, within their differential diagnosis, they frequently rejected the need for definitive investigations like a lumbar puncture. This demonstrates that while an LLM can successfully pattern-match a patient’s symptoms to a textbook differential diagnosis, it does not necessarily translate in a safe management plan. The models lack the clinical heuristics required to weigh highly conditional management pathways and balance the catastrophic cost of missing a low likelihood but possibly fatal diagnosis against the minor morbidity of a diagnostic test.

This inverse relationship between information density and patient safety demonstrates that current LLMs lack the deductive caution central to medical practice^2,3^. To safely integrate these tools into healthcare settings, future models must be equipped with explicit algorithmic guardrails. Rather than optimising solely for probabilistic efficiency, their underlying logic must be recalibrated to mirror human risk-mitigation, ensuring that the clinical imperative to rule out ‘must-not-miss’ pathology always supersedes simple statistical likelihood.

The models also demonstrated some consistent blind spots which persisted regardless of information completeness. In presentations of idiopathic intracranial hypertension featuring symptoms of raised intracranial pressure, even when LLMs achieved 100% accuracy in diagnosing it, they failed to reliably identify the need for lumbar puncture despite it being both the definitive diagnostic standard and an acute therapeutic, potentially sight-saving, intervention. The inability to safely manage uncertainty also produced demographic biases which resulted in female patients, particularly those aged 30 to 50, being disproportionately advised to self-manage or have routine follow-up for life- or sight-threatening emergency conditions.

Exposing and correcting these sorts of lethal reasoning failures requires moving beyond traditional evaluation metrics. By stress-testing models across nearly 3 million words of dynamic patient dialogue, this methodology allowed us to evaluate 14,000 discrete medical recommendations made by LLMs. This scale and precision are entirely unattainable through manual human review, and it is only at this large scale that subtle demographic biases and dangerous probabilistic heuristics of frontier models can become visible.

This methodology enables rapid generation of thousands of diverse patient encounters at low cost, and by mapping every model decision back to formal diagnostic criteria in simulation, it can transition LLM evaluation from simple accuracy scoring to granular, explainable audits of clinical reasoning. We can map exact points of algorithmic failure and identify vulnerable patient profiles before unproven models are ever deployed into live clinical environments.

The computational efficiency of this approach also enables potential counterfactual analysis. If an LLM fails, we can rapidly generate thousands of micro-variations of that specific encounter to pinpoint the clinical trigger(s) for the error. While piloted in headache medicine, this simulation methodology can be easily applied in other clinical domains^28^ to enable testing across a large spectrum of human communication and clinical complexity, rather than just a few of the most typical textbook presentations.

While simulating real clinical practice presents inherent challenges, we implemented proactive measures to mitigate these limitations. First, artificial transcripts do not perfectly encapsulate all possible unpredictable ways in which real patients may report symptoms. We addressed this by explicitly modelling seven dimensions of patient persona and injecting realistic linguistic noise into the histories with use of an LLM set to a high temperature setting. Second, written transcripts cannot capture non-verbal cues, such as a patient’s tone of voice or physical distress, which inherently influence human assessments. While the impact of non-verbal communication requires future characterisation, it does not invalidate the reasoning failures identified in the LLMs’ text-processing capabilities. Third, our current simulation focuses on single primary diagnoses, whereas real patients frequently present with multimorbidity. We plan to expand this platform to simulate mixed phenotypes, to assess how LLMs navigate conflicting information and complex overlapping management pathways. Finally, commercial LLMs undergo continuous and often opaque updates; a model deemed safe today may behave differently tomorrow. However, because our computationally lightweight approach can rapidly generate and analyse thousands of cases on demand, it provides an ideal engine for continuous, automated post-deployment monitoring, ensuring any degradation in safety can be identified promptly.

Safe integration of AI in healthcare requires confronting the limitations of LLM probabilistic reasoning^29,30^. This work highlights three components of this. First, model selection is highly consequential. The stark performance discrepancies observed here between leading LLMs highlights that these tools cannot be treated interchangeably. Rigorous evidence and total transparency should be a mandatory prerequisite for use of LLMs in any healthcare setting, adhering to the same standards as pharmaceuticals^26^. Second, LLMs possess particularly dangerous blind spots when faced with incomplete information. They unsafely equate an absence of evidence with an absence of disease. Where human clinicians naturally adopt caution to rule out ‘must-not-miss’ catastrophic outcomes, these probabilistic systems actively de-escalated care and dismissed life-saving investigations precisely when a patient’s history was most ambiguous. Third, exposing and correcting these lethal reasoning failures requires moving beyond traditional evaluation metrics. The high-throughput simulation methodology introduced here provides a novel approach for clinical AI evaluation. Only by stress-testing models across thousands of diverse, dynamically generated, non-idealised scenarios, can we hope to map exact points of failure and to engineer the algorithmic guardrails required to prioritise patient safety before ever exposing real patients to these risks.

## ONLINE METHODS

### Case generation

We developed a patient simulation engine to generate a highly controlled dataset of 1,000 synthetic transcripts of doctor-patient encounters, encompassing 33 primary and secondary headache, and facial pain diagnoses. Diagnoses were selected based on the International Classification of Headache Disorders, 3rd edition (ICHD-3)^21^.

We enforced a balanced distribution across all 33 syndromes to evaluate clinical reasoning equally across common primary headaches (e.g. migraine with aura), rarer conditions (e.g. SUNCT), and secondary medical emergencies (e.g. brain tumours or giant cell arteritis). We intentionally did not attempt to closely model patient demographic (age, sex) or diagnostic prevalence based on natural epidemiological frequencies as relying on ‘realistic’ epidemiological distributions inherently biases models toward the most statistically common presentations.

To preserve the integrity of this simulation engine as a rigorous, future-proof evaluation tool for frontier medical AI, we do not publicly release the precise generative codebase or the exact probability distributions used to define the simulated cohorts. Making these granular parameters openly accessible in the training corpus of future LLMs would risk data contamination, allowing models to inadvertently memorise or overfit to our evaluation distributions. Instead, we detail the comprehensive logical architecture and principles of the methodology required to reproduce this simulation approach independently.

For each case, we first sampled a precise set of underlying headache phenotype facts. The sampling logic was derived directly from ICHD-3 criteria, which dictated the essential phenotypic features required to ‘rule in’ a given diagnosis. Where formal ICHD-3 criteria lacked complete clinical context, supplementary pathophysiological constraints were applied (e.g. restricting giant cell arteritis to simulated patients with age >50 years). To construct a complete clinical case, we populated these required fields based on the ICHD-3 constraints, while filling any remaining, non-constrained clinical fields (e.g. exact pain score out of 10, specific time of day of onset) by sampling uniformly from predefined, clinically plausible ranges. By iterating through all possible features of a headache phenotype and sampling unique combinations of values for every case, we generated a dataset that strictly satisfied diagnostic criteria while covering the full spectrum of possible typical and atypical presentations. All logic matrices and medical evidence were constructed, reviewed and validated by a UK-registered specialist neurologist (SDA). This process yielded a set of deterministic ground-truth labels for every generated case, allowing every clinical variable within a simulation to be mapped directly back to the specific diagnostic rule it satisfied.

### Transcript generation

These deterministic clinical facts were transformed into naturalistic conversational transcripts using a targeted LLM (GPT-OSS 20B^22^; temperature = 0.7 to maximise linguistic variance). To isolate the impact of patient-side communication on clinical reasoning, we held the doctor’s questioning constant across all encounters. We simulated a ‘doctor’ adhering to a strictly pre-defined clinical script. Every transcript began with identical open-ended questions (e.g. *“What is the main symptom you are seeing the doctor for today?“* and *“Could you please tell me more about your [headache/face pain] in your own words?“*), followed by a narrowing sequence of targeted, closed-ended questions (e.g. a query regarding headache duration used identical phrasing in every case).

To capture the fragmented, idiosyncratic reality of real-world communication, patient responses were systematically modulated by sampling a unique, seven-dimensional persona for each case:

1. Verbosity: Defined informational density across three tiers: extreme brevity (fragments/single words, maximum 15 words per turn), normal length (standard sentences), and verbose (rambling context, up to 120 words per turn).
2. Literacy: Ranged across five tiers, from basic nouns/verbs to a medical expert tier using precise anatomical and medical terminology. Intermediate tiers included low-average, average, and highly articulate non-medical academic language.
3. English Proficiency: Scaled from broken/learner English (dropped articles, mixed tenses) through functional non-native phrasing (idiomatic errors) to native fluency.
4. Memory Reliability: Ranged from unreliable (uncertain timelines), to fuzzy (rounding approximations), to sharp (precise recall of dates and dosages).
5. Focus: Dictated conversational relevance, defined as tangential (frequent drifting), conversational (occasional filler), or laser-focused (efficient, zero unrelated information).
6. Disclosure: Reflected willingness to share, ranging from guarded (requiring explicit prompting), through passive neutrality, to oversharing (volunteering extra information unprompted).
7. Framing: Captured the subjective perception of pain, operationalised as stoic (minimising suffering), objective (matter-of-fact), or catastrophising (highly anxious language).

Sampling across these seven domains gave rise to 3,645 possible unique patient personas.

For each doctor enquiry, the generative model synthesised an answer by drawing the specific clinical content from the phenotype facts, while simultaneously adopting the sampled persona to dictate the tone, style, and linguistic structure for the response. An automated blinded secondary LLM verification step ensured the generated dialogue strictly adhered to the hardcoded phenotype facts without introducing ‘hallucinated’ clinical features before outputting the patient answer.

To explicitly test how models navigate the presence of incomplete information, we systematically varied the completeness of the clinical history. For each transcript, the available clinical facts were restricted *a priori* to one of five predefined ‘sampling rates’ (20%, 40%, 60%, 80%, or 100%). The assigned sampling rate solely dictated *which* of the standardised doctor questions were asked and which patient phenotype facts were available in their answers. To ensure clinical coherence across different headache types, we did not sample individual data points in isolation; instead, we grouped related features into logical sampling units.

For example, a single ‘unit’ might consist of all questions related to aura, or the single question defining headache duration. By calculating the sampling rate based on these units rather than a raw count of facts, we maintained a consistent level of ‘missingness’ across different headache types, regardless of whether a specific diagnosis required four or six criteria to satisfy. A 100% sampling rate included every unit required for an ICHD-3 diagnosis; a 20% rate included only one-fifth of those available units, simulating a highly incomplete clinical history. Each of 1,000 cases was randomly assigned to one of the five information sampling levels (20% to 100%), ensuring the final dataset represented a broad spectrum of clinical completeness across the various headache phenotypes. At the end of transcripts, a statement was added indicating that patients had a normal neurological examination.

No two cases were identical, but all cases adhered to the relevant diagnostic criteria. Variation occurred both in the underlying clinical phenotype, the narrative delivery and how complete the information available in a transcript was.

The simulation engine produced a primary evaluation dataset of 1,000 unique consultation transcripts, comprising 68,690 lines of conversational dialogue and 1,435,631 words.

### LLM evaluation protocol

We evaluated two frontier LLMs: GPT-5.2 (OpenAI’s state-of-the-art reasoning model at the time of running the experiment) and GPT-5-mini (the smaller architecture reportedly powering public-facing health tools^2^). Models were evaluated using a zero-shot prompting strategy to prevent in-context learning. Hyperparameters were set to standard defaults and internal reasoning protocols were configured to their ‘high’ settings to elicit maximum performance.

For each transcript, the models were tasked with: (1) providing a comprehensive prioritised differential diagnosis; (2) estimating the percentage likelihood for each provided differential; (3) making specific management decisions regarding diagnostic procedures (CT head, MRI head, lumbar puncture) and pharmacology (indomethacin, codeine); and (4) recommending an appropriate timeline for clinical follow-up based solely on the provided history.

To mitigate framing bias, the phrasing of management questions was randomised per case (e.g. asking whether to “request” versus “cancel” a scan which had already been requested; “start” versus “stop” a medication). For investigational and procedural questions, the models were strictly required to select one of three outputs: *“Yes“*, *“No“*, or *“Inappropriate to decide from current information“*. Treatment questions required binary Yes/No answers.

For follow-up triage, models selected between: *“Immediate“* (emergency department attendance), *“Urgent“* (follow-up within 24–48 hours), *“Routine“* (follow-up within 1–6 months), or *“Self-management“* (no scheduled clinician follow-up necessary). All questions were presented to the model independently to ensure that preceding enquiries did not implicitly anchor or bias subsequent reasoning. Transcripts and prompts were submitted to, and LLM answers retrieved from, OpenAI’s batch application programming interface.

### Data Processing and Statistical Analysis

Because both GPT-5.2 and GPT-5-mini were evaluated independently against the entire dataset, the total inferential burden across the study encompassed 137,380 lines of dialogue and 2,871,262 words. From this vast conversational corpus, the automated parsing pipeline extracted and statistically analysed exactly 14,000 distinct clinical determinations (encompassing diagnostic accuracy, procedural interventions, pharmacological prescribing, and triage acuity).

Data processing and statistical analyses were conducted using Python (v3.12), including use of the pandas, scipy, seaborn, matplotlib and statsmodels libraries. LLM text outputs were automatically parsed, standardised via exact string matching to remove whitespace artifacts, and compared against the deterministic ground-truth labels.

Diagnostic accuracy was defined as the successful inclusion of the ground-truth diagnosis anywhere within the model’s generated differential list. The classification of indomethacin-responsive conditions, along with the supporting clinical evidence for these determinations, is detailed in Supplementary Table 1.

For the assessment of follow-up safety, an unsafe triage was defined as a model recommending either *“Routine“* or *“Self-management“* for any patient with a life-threatening or vision-threatening emergency diagnosis; namely any of acute angle-closure glaucoma, giant cell arteritis, brain tumours, idiopathic intracranial hypertension with signs of raised intracranial pressure, meningitis, reversible cerebral vasoconstriction syndrome, or subarachnoid haemorrhage. For demographic analyses, simulated ages were grouped into categorical buckets (<30, 30–50, 50–70, >70 years) and time since symptom onset was standardised to days to calculate acuity cohorts (e.g. <14 days).

Statistical methodologies were tailored to the experimental design. Because both models (GPT-5.2 and GPT-5-mini) were evaluated on the identical set of 1,000 patient transcripts, their outputs constitute paired, dependent data. Therefore, all inter-model statistical comparisons (e.g. comparing the accuracy or investigation rates of GPT-5.2 versus GPT-5-mini) were conducted using McNemar’s test for paired nominal data.

Conversely, comparisons made *within* a single model across independent case variables, such as comparing performance across different sampling rates (e.g., 20% vs. 100% information density), patient biological sex, or age demographics, were evaluated using Pearson’s Chi-square test of independence.

To provide robust estimates of variance for all reported proportions, 95% confidence intervals (CIs) were calculated using a non-parametric bootstrapping method. For each target proportion, the sample data was resampled with replacement for 10,000 iterations to generate a distribution of sample proportions. The 95% CI was defined by the 2.5th and 97.5th percentiles of this bootstrapped distribution. Statistical significance was defined *a priori* as *P* < 0.05 (two-tailed). All reported metrics in the main text are presented as the mean proportion accompanied by its bootstrapped 95% CI.

To quantify the breadth of diagnostic reasoning, we calculated the number of unique diagnoses provided by each model. We assessed the relationship between the sampling rates of 20%–100% and differential diagnosis length using the Kruskal-Wallis H test to detect differences in distributions across tiers.

## Data and code availability

The results, processed data and analysis code supporting the findings of this study will be made freely available upon publication at https://github.com/stepdaug. However, the raw simulated patient transcripts and the exact probability distributions used to define the simulated cohorts are not publicly available. These restrictions are in place to preserve the integrity of the simulation engine as a rigorous, future-proof evaluation tool for frontier medical AI. Public release of these datasets would risk contamination of future model training sets, thereby compromising the tool’s utility as a neutral benchmark. Requests for access to the transcripts for non-commercial research purposes may be considered by the authors upon request.

## Acknowledgements

SDA is supported by a UK National Institute for Health and Care Research (NIHR) Clinical Lectureship and acknowledges infrastructure support for this research from the NIHR Imperial Biomedical Research Centre (BRC).

GS, NIHR Advanced Fellow, NIHR302971 is funded by the NIHR for this research project. The views expressed in this publication are those of the author and not necessarily those of the NIHR, NHS or the UK Department of Health and Social Care

The authors also acknowledge MRC equipment support via the UK Dementia Research Institute and are grateful to Payam Barnaghi for assistance in accessing this.

## Author Contributions

SDA conception, design, data analysis, manuscript first draft and review. GS design and manuscript review.

