## Supplementary information for "Medical errors in large language models revealed using 1,000 synthetic clinical transcripts"

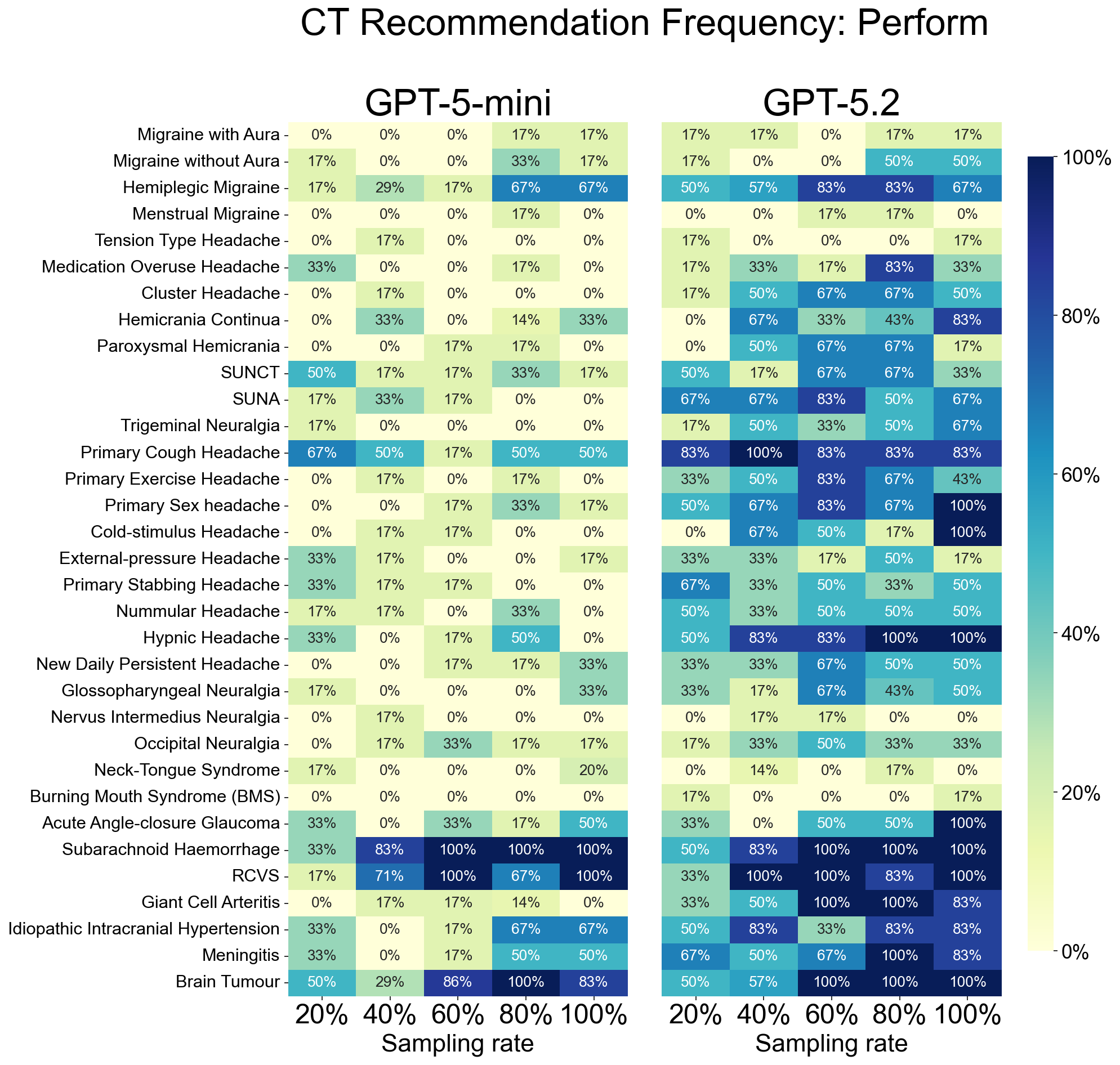

**Supplementary Figure 1 | Frequency of CT head scan recommendation (Perform).** Heatmaps displaying the frequency with which LLMs recommended a CT head scan should be performed, stratified by clinical history completeness (sampling rate; 20% to 100%). Colour intensity represents the proportion of encounters where the model recommended performing a scan (dark blue = 100%; light yellow = 0%). GPT-5-mini (left) and GPT-5.2 (right) are displayed across 33 distinct headache and facial pain syndromes. Syndromes are ordered from common primary headaches to urgent secondary causes. SUNCT - Short-lasting unilateral neuralgiform headache with conjunctival injection and tearing, SUNA - Short-lasting Unilateral neuralgiform headache with cranial autonomic symptoms, RCVS - Reversible Cerebral Vasoconstriction Syndrome.

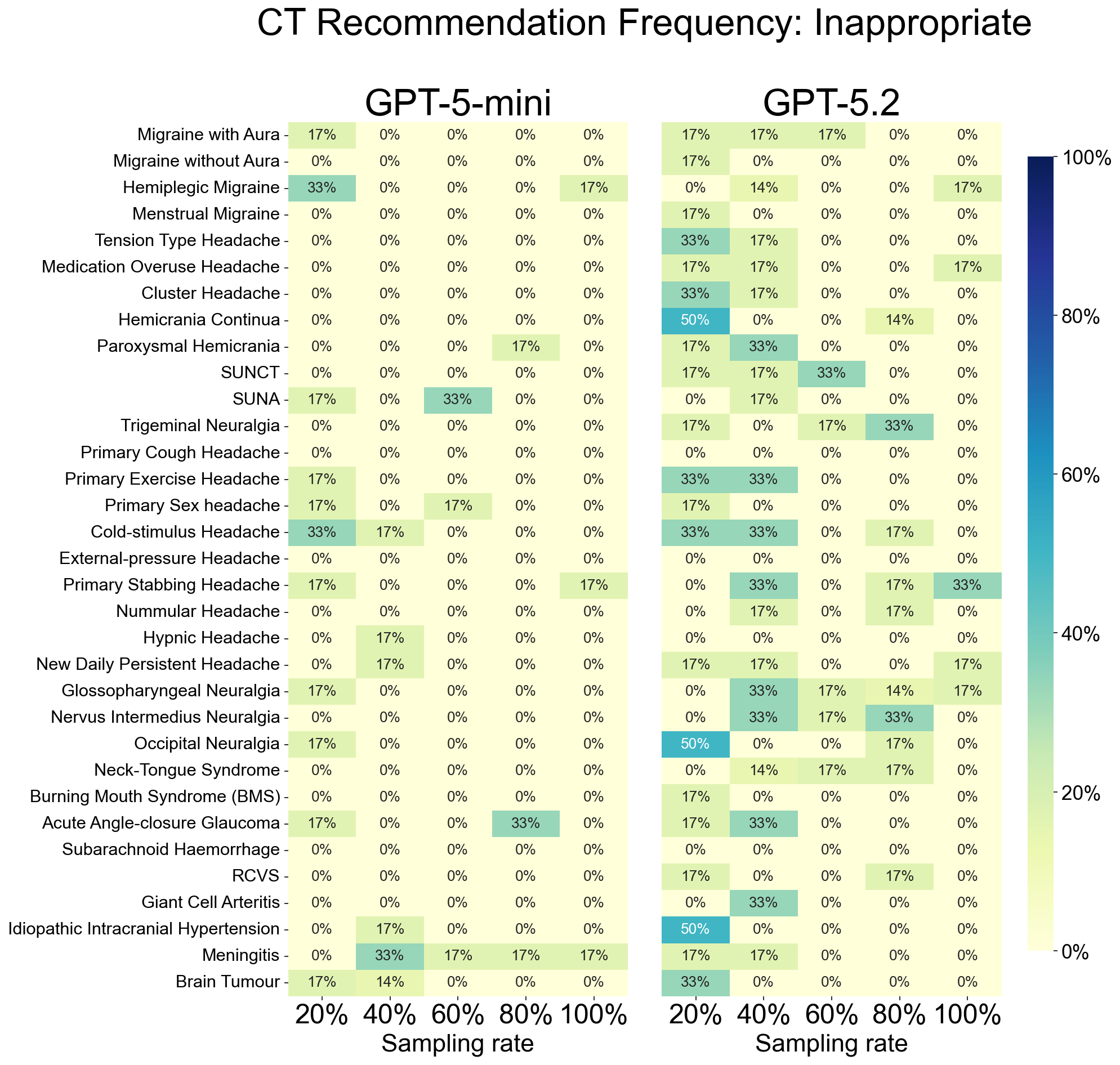

**Supplementary Figure 2 | Frequency of CT head scan recommendation (Inappropriate).** Heatmaps displaying the frequency with which LLMs classified a CT head scan as ‘inappropriate to decide’ from current information for the provided clinical vignette. Colour intensity represents the proportion of encounters (dark blue = 100%; light yellow = 0%). GPT-5-mini (left) and GPT-5.2 (right) recommendations are displayed across 33 headache and facial pain syndromes.

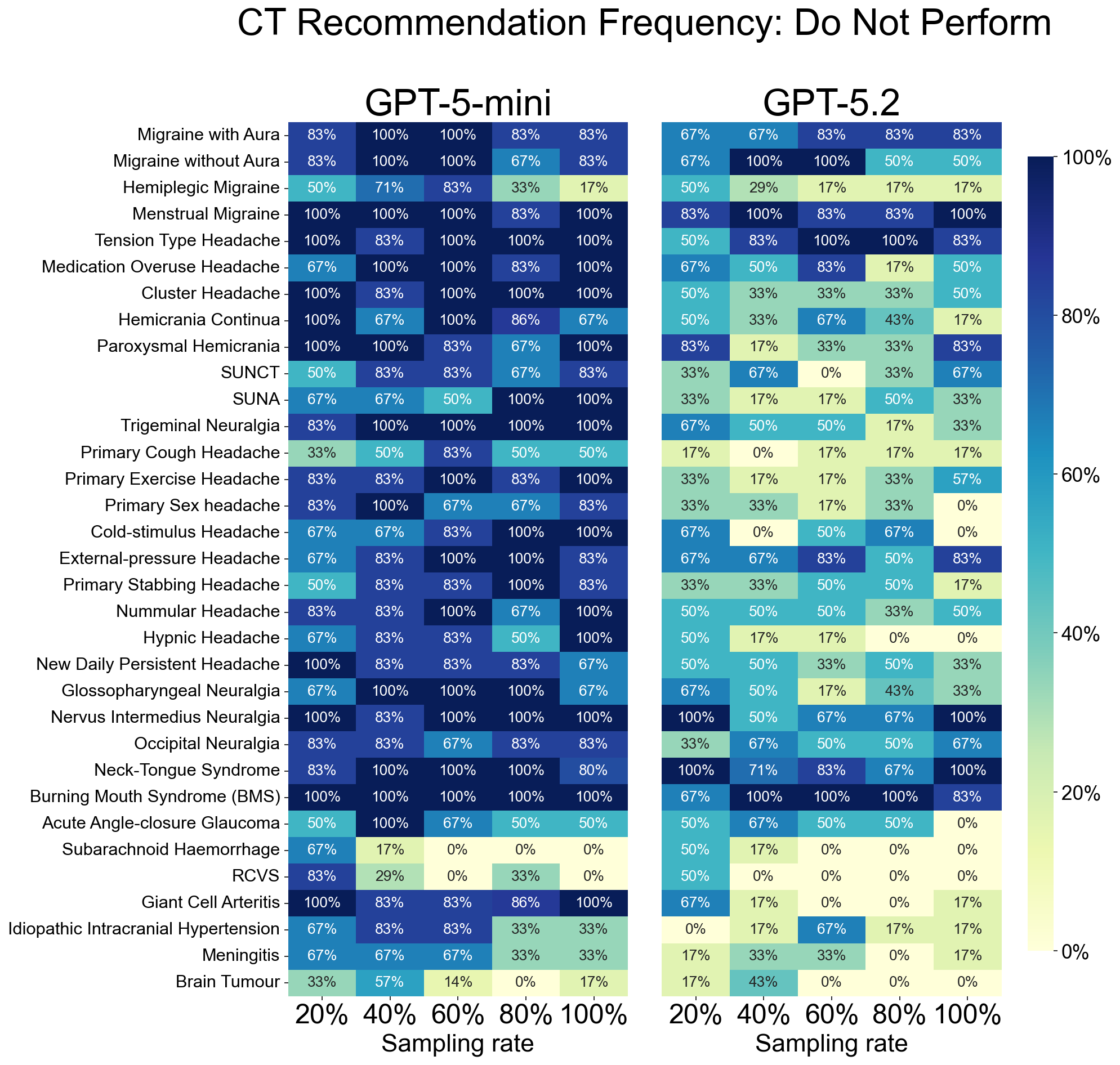

**Supplementary Figure 3 | Frequency of CT head scan recommendation (Do Not Perform).** Heatmaps displaying the frequency with which LLMs explicitly recommended against performing a CT head scan. Colour intensity represents the proportion of encounters (dark blue = 100%; light yellow = 0%). GPT-5-mini (left) and GPT-5.2 (right) recommendations are displayed across 33 headache and facial pain syndromes.

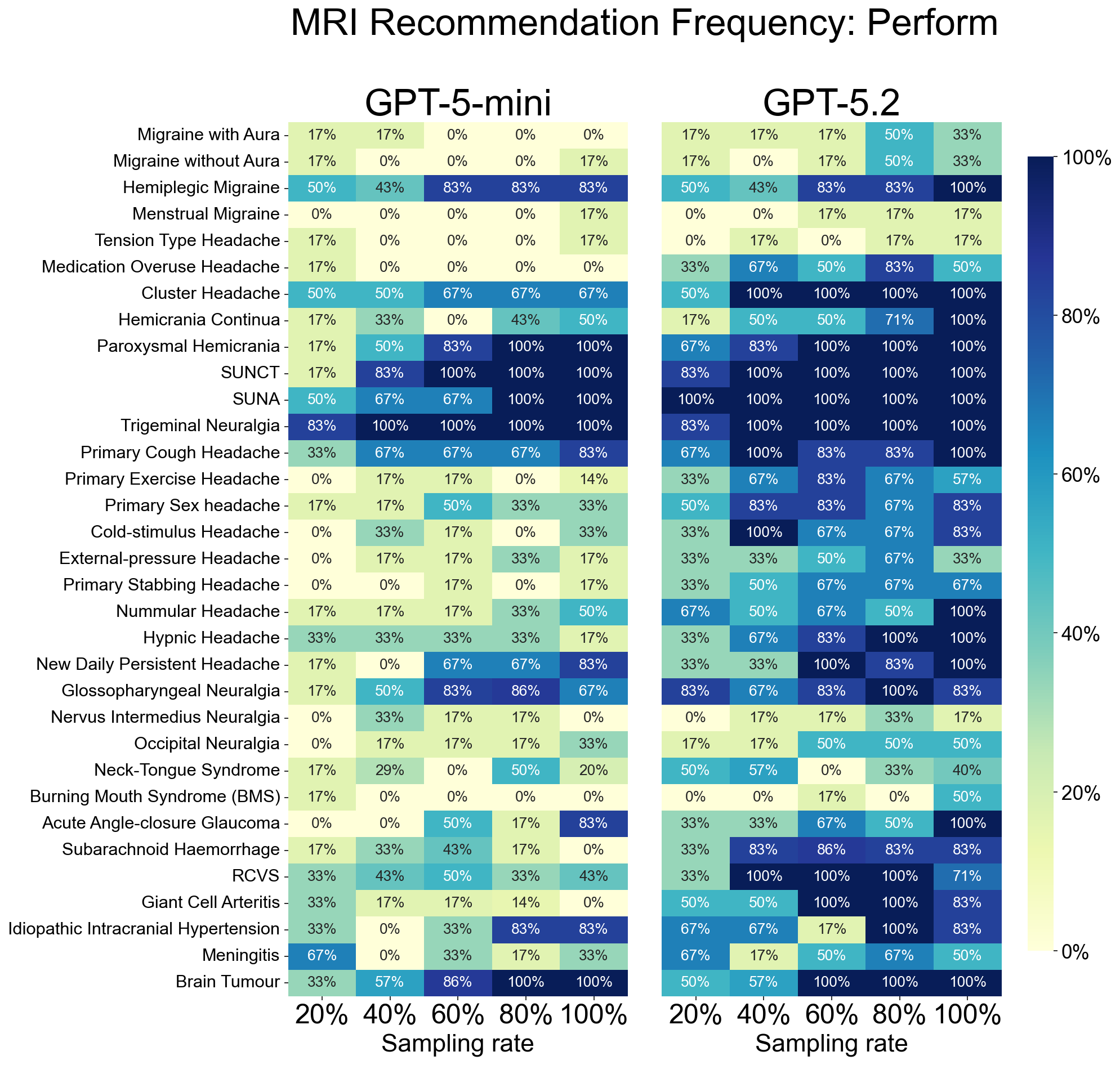

**Supplementary Figure 4 | Frequency of MRI head scan recommendation (Perform).** Heatmaps displaying the frequency with which LLMs recommended an MRI head scan, stratified by clinical history completeness. Colour intensity represents the proportion of encounters where the model recommended performing a scan (dark blue = 100%; light yellow = 0%). GPT-5-mini (left) and GPT-5.2 (right) are displayed across 33 headache and facial pain syndromes.

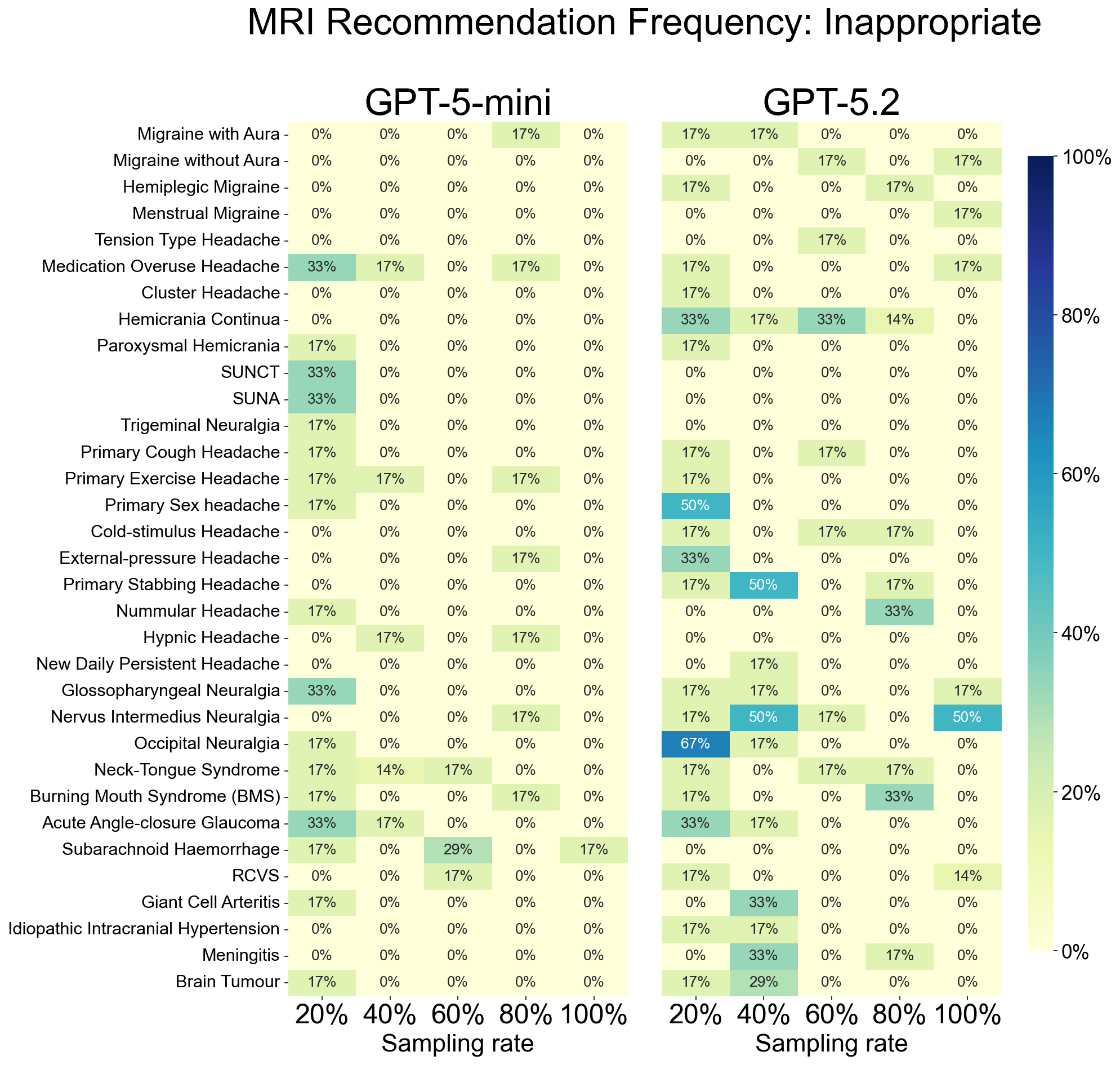

**Supplementary Figure 5 | Frequency of MRI head scan recommendation (Inappropriate).** Heatmaps displaying the frequency with which LLMs classified an MRI head scan as ‘inappropriate to decide’ from current information for the provided clinical vignette. Colour intensity represents the proportion of encounters (dark blue = 100%; light yellow = 0%). GPT-5-mini (left) and GPT-5.2 (right) are displayed across 33 headache and facial pain syndromes.

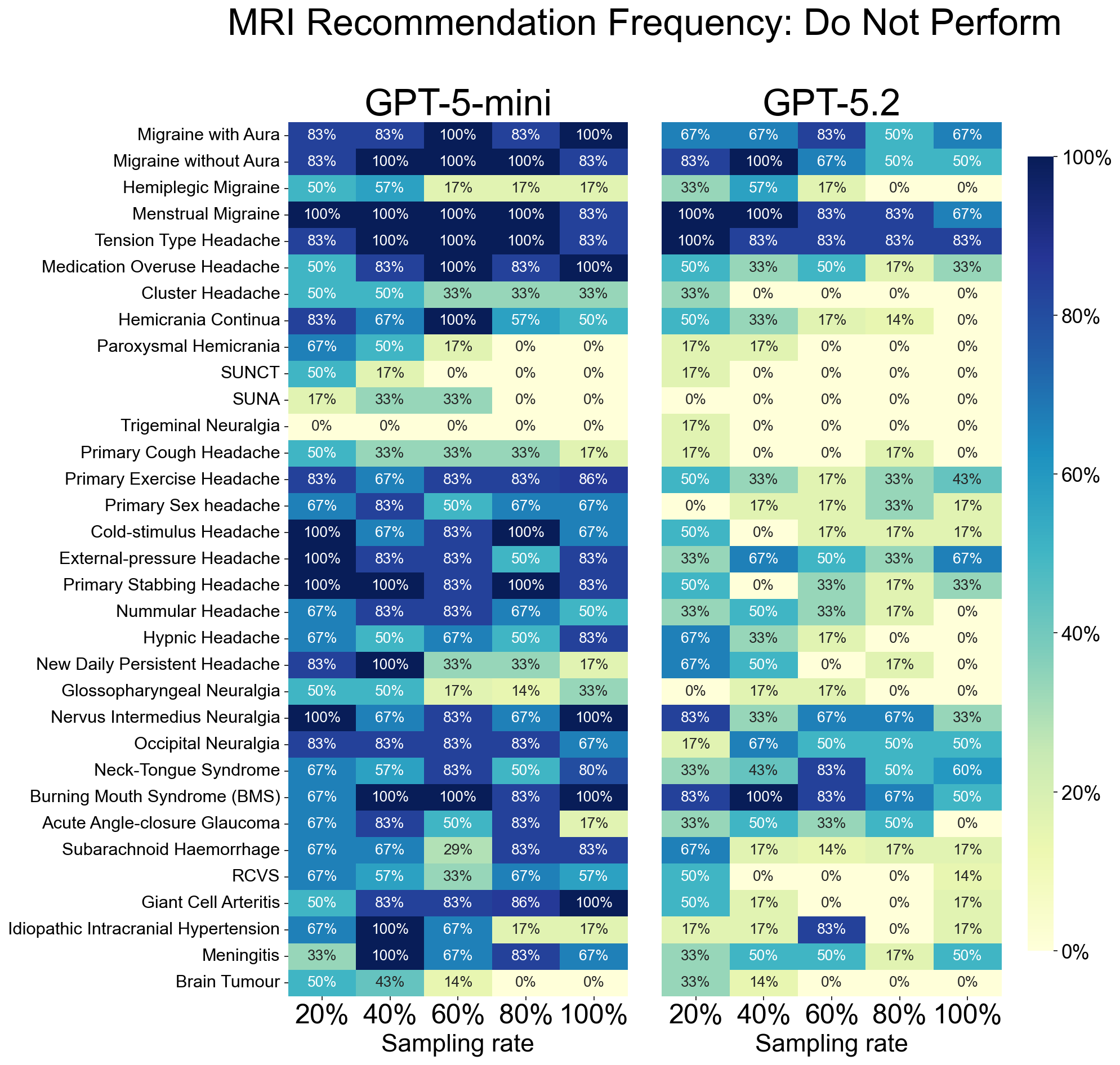

**Supplementary Figure 6 | Frequency of MRI head scan recommendation (Do Not Perform).** Heatmaps displaying the frequency with which LLMs explicitly recommended against performing an MRI head scan. Colour intensity represents the proportion of encounters (dark blue = 100%; light yellow = 0%). GPT-5-mini (left) and GPT-5.2 (right) are displayed across 33 headache and facial pain syndromes.

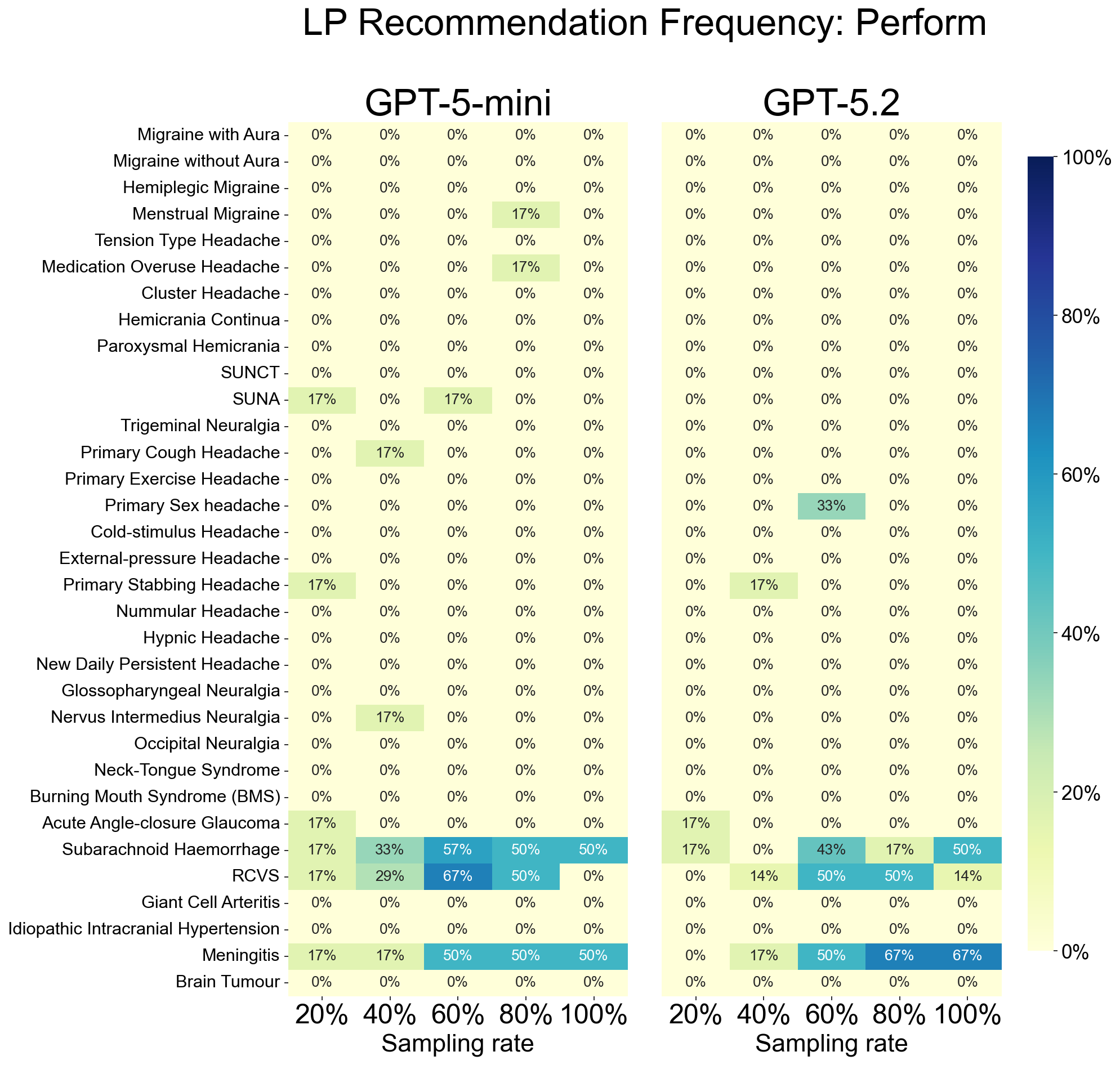

**Supplementary Figure 7 | Frequency of Lumbar Puncture recommendation (Perform).** Heatmaps displaying the frequency with which LLMs recommended a lumbar puncture, stratified by clinical history completeness. Colour intensity represents the proportion of encounters where the model recommended performing a procedure (dark blue = 100%; light yellow = 0%). GPT-5-mini (left) and GPT-5.2 (right) are displayed across 33 headache and facial pain syndromes.

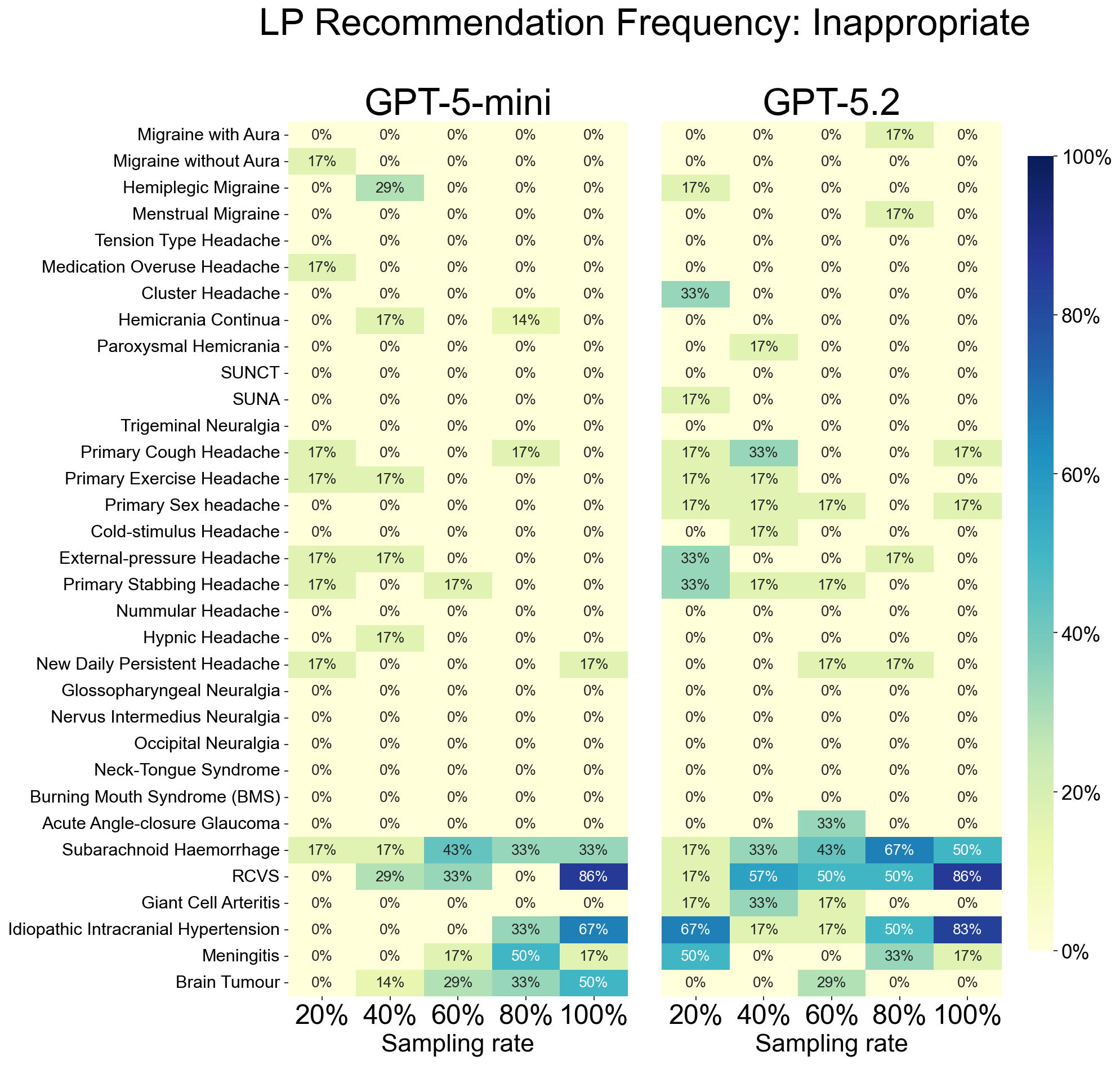

**Supplementary Figure 8 | Frequency of Lumbar Puncture recommendation (Inappropriate).** Heatmaps displaying the frequency with which LLMs classified a lumbar puncture as ‘inappropriate to decide’ from current information for the provided clinical vignette. Colour intensity represents the proportion of encounters (dark blue = 100%; light yellow = 0%). GPT-5-mini (left) and GPT-5.2 (right) are displayed across 33 headache and facial pain syndromes.

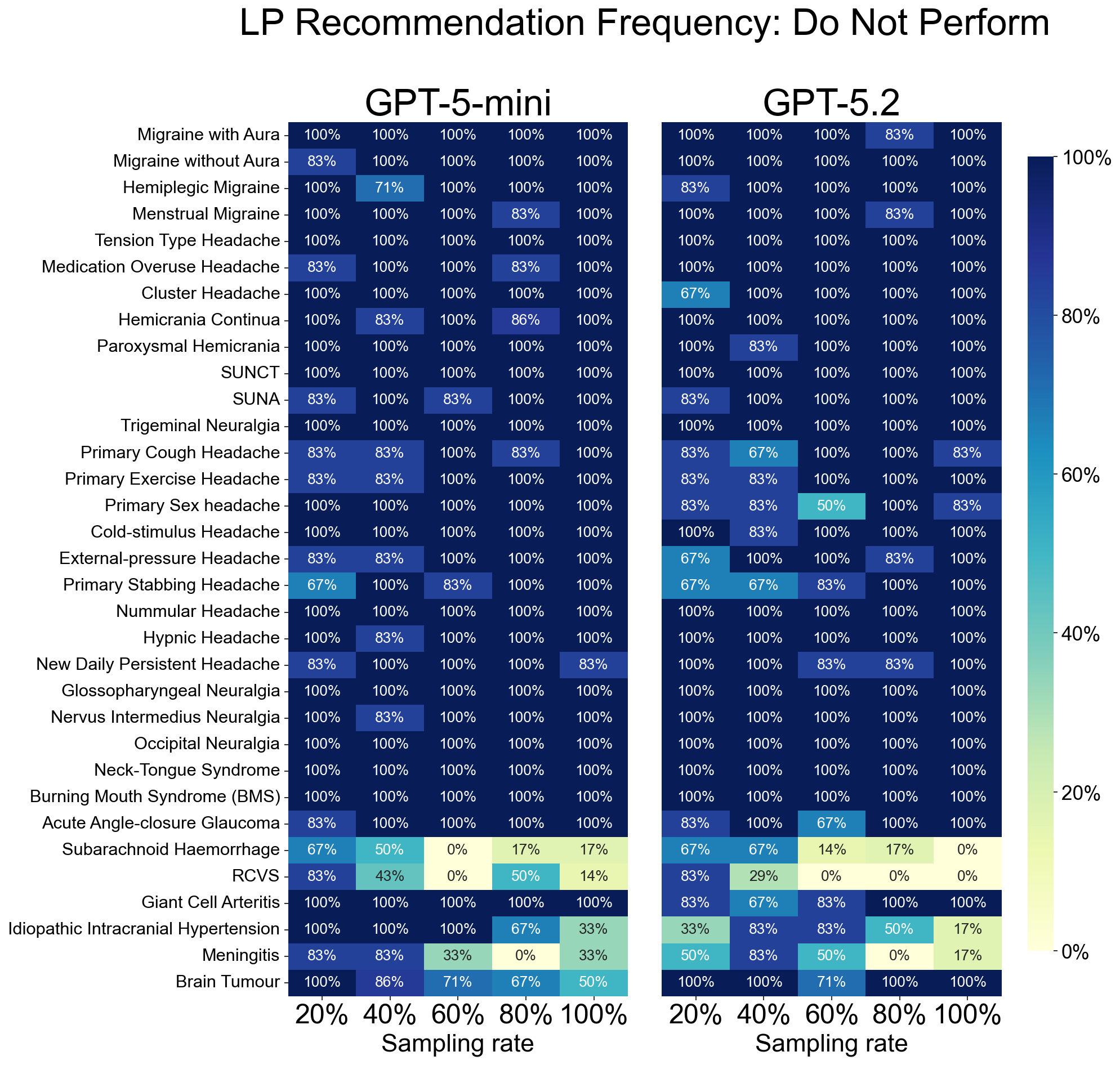

**Supplementary Figure 9 | Frequency of Lumbar Puncture recommendation (Do Not Perform).** Heatmaps displaying the frequency with which LLMs explicitly recommended against performing a lumbar puncture. Colour intensity represents the proportion of encounters (dark blue = 100%; light yellow = 0%). GPT-5-mini (left) and GPT-5.2 (right) are displayed across 33 headache and facial pain syndromes.

**Supplemental Table 1 - Ground truth used for indomethacin management decisions**

| **Diagnosis** | **Indomethacin always appropriate** | **Indomethacin never appropriate** | **Indomethacin sometimes appropriate** | **References** |
| --- | --- | --- | --- | --- |
| Migraine with Aura |  | **Yes** |  | ^1–5^ |
| Migraine without Aura |  | **Yes** |  | ^1–5^ |
| Tension Type Headache |  | **Yes** |  | ^1–5^ |
| Medication Overuse Headache |  | **Yes** |  | ^1–5^ |
| Cluster Headache |  |  | **Yes** (rare case reports) | ^2,4,5^ |
| Hemicrania Continua | **Yes** |  |  | ^4–6^ |
| Paroxysmal Hemicrania | **Yes** |  |  | ^4–6^ |
| SUNCT |  | **Yes** |  | ^4,6^ |
| SUNA |  | **Yes** |  | ^4,6^ |
| Trigeminal Neuralgia |  | **Yes** |  | ^2,4,5^ |
| Giant Cell Arteritis |  | **Yes** |  | ^2,4,5^ |
| Hemiplegic Migraine |  | **Yes** |  | ^2,4,5^ |
| Menstrual Migraine |  | **Yes** |  | ^2,4,5^ |
| Primary Cough Headache |  |  | **Yes** | ^1,2,4,5^ |
| Primary Exercise Headache |  |  | **Yes** | ^1,2,4,5^ |
| Primary Headache Associated with Sexual Activity |  |  | **Yes** | ^1,2,4,5^ |
| Cold-stimulus Headache |  | **Yes** |  | ^2,4,5^ |
| External-pressure Headache |  | **Yes** |  | ^2,4,5^ |
| Primary Stabbing Headache |  |  | **Yes** | ^2,4,5^ |
| Nummular Headache |  |  | **Yes** (rare case reports) | ^2,4,5^ |
| Hypnic Headache |  |  | **Yes** | ^1,2,4,5^ |
| New Daily Persistent Headache |  | **Yes** |  | ^2,4,5^ |
| Subarachnoid Haemorrhage |  | **Yes** |  | ^2,4,5^ |
| Reversible Cerebral Vasoconstriction Syndrome |  | **Yes** |  | ^2,4,5^ |
| Glossopharyngeal Neuralgia |  | **Yes** |  | ^2,4,5^ |
| Nervus Intermedius Neuralgia |  | **Yes** |  | ^2,4,5^ |
| Occipital Neuralgia |  | **Yes** |  | ^2,4,5^ |
| Neck-Tongue Syndrome |  | **Yes** |  | ^2,4,5^ |
| Burning Mouth Syndrome |  | **Yes** |  | ^2,4,5^ |
| Headache due to Brain Tumour |  | **Yes** |  | ^2,4,5^ |
| Headache attributed to Acute Angle-closure Glaucoma |  | **Yes** |  | ^2,4,5^ |
| Idiopathic Intracranial Hypertension |  | **Yes** |  | ^2,4,5^ |
| Headache attributed to Meningitis |  | **Yes** |  | ^2,4,5^ |
